# Bronchial gene expression clustering in COPD identifies a subgroup of patients with higher level of bronchial T-cells and accelerated lung function decline

**DOI:** 10.1101/2025.07.21.25331922

**Authors:** Rui Marçalo, Alen Faiz, Orestes A. Carpaij, Tatiana Karp, Jeunard Boekhoudt, Avrum Spira, Judith M. Vonk, Wim Timens, Hubertus A. M. Kerstjens, Gaik W. Tew, Michele A. Grimbaldeston, Margaret Neighbors, Pieter S. Hiemstra, Lisette I. Z. Kunz, Stephen Lam, Victor Guryev, Alda Marques, Gabriela R. Moura, Katrina Steiling, Corry-Anke Brandsma, Maarten van den Berge

## Abstract

Chronic obstructive pulmonary disease (COPD) is a heterogeneous disease with varying degrees of airway wall thickening, chronic bronchitis, and emphysema. A better understanding of the underlying pathology is needed to improve the personalized treatment of the disease and identify new therapeutic targets.

Available data from 56 COPD patients included in the GLUCOLD study were used (61.1±7.7 years, 89% male, and FEV_1_ of 62.5±8.9% predicted). Clinical characterization was performed including bronchoscopy and collection of bronchial biopsies at baseline. RNA from bronchial biopsies was sequenced and used for unsupervised clustering, using a 98 COPD gene signature previously identified in bronchial brushes comparing patients with COPD to non-COPD controls. Next, we assessed differences in the clinical expression of COPD, lung function decline, inflammatory cell counts, and gene expression between clusters. Validation was performed in an independent dataset.

We identified two clusters: CAGE1 (n=39) and CAGE2 (n=17). CAGE2 patients had higher percentage of sputum lymphocytes, and more CD4^+^ and CD8^+^ T-cell counts in their bronchial biopsies. In addition, their FEV_1_ improved less in response to 30-months treatment with inhaled corticosteroids (ICS) (change in FEV_1_- CAGE1: +24.4mL; CAGE2: −29.1mL; p-value=0.048), and they experienced a faster decline in their lung function follow-up (CAGE1: −44.0mL/year; CAGE2: −69.9mL/year; p-value=0.002). Gene expression analysis showed more activation of T- and B-cell immune responses in CAGE2.

We identified a new COPD endotype, CAGE2, characterized by ICS unresponsiveness and faster lung function decline. Additionally, we show a different pathobiology in CAGE2 with more activation of T- and B-cell immune responses.

## Introduction

Chronic obstructive pulmonary disease (COPD) is a complex condition characterized by diverse manifestations, such as airway wall thickening, chronic bronchitis and emphysema, contributing to chronic airflow obstruction. There are currently no pharmacological treatments available that prevent the progressive decline in lung function associated with COPD. A deeper understanding of the underlying pathology is essential to refine personalized treatment approaches and uncover novel therapeutic targets.

Genome-wide gene expression profiling offers a promising avenue to categorize subgroups of patients with obstructive pulmonary diseases based on their unique pathobiological mechanisms, a concept known as endotyping(1). This approach provides valuable insight into the specific disease components that may be amenable for intervention in individual patients, thereby potentially improving their precision and effectiveness. Previous studies have employed unsupervised gene expression cluster analyses on various biological samples such as blood, sputum, and bronchial biopsies(2–7). As an example, three asthma clusters were identified based on sputum gene expression profiling. These asthma clusters were shown to reflect different underlying pathobiological mechanisms that could be related to distinct inflammatory profiles (first cluster was characterized by high eosinophils levels, second cluster was characterized by high neutrophils levels, whereas the third cluster was characterized by high macrophages levels) and were linked to distinct clinical features (first cluster was characterized by poorer asthma control, whereas third cluster was characterized by better lung function)(3). In another study, using blood microarray gene expression data in 141 patients with COPD and 88 smoker controls, unsupervised clustering revealed four distinct endotypes(7). One of the endotypes was associated with severe lung function impairment, respiratory symptoms, and emphysema, whereas another endotype exhibited preserved lung function and less emphysema. Despite these advances, no studies have applied this approach directly to the airway wall in COPD, closer to the site where the disease actually occurs.

We have previously showed the expression of 98 genes to be associated with presence of COPD. Although statistically significant, there was still considerable variability in the expression of these genes within COPD patients, likely reflecting disease heterogeneity. To better understand the different endotypes, we performed unsupervised clustering of available RNA-sequencing data obtained from bronchial biopsies of well-characterized patients with COPD from the Groningen and Leiden Universities Study of Corticosteroids in Obstructive Lung Disease (GLUCOLD) study, using this 98 gene signature(8). We correlated the resulting clusters with physiological and histopathological parameters to identify clinically relevant COPD endotypes.

## Methods

### Study design

This study is a secondary analysis of patients who participated in the GLUCOLD study (ClinicalTrials.gov registration number: NCT00158847)(9). GLUCOLD was a randomized, double-blind, placebo-controlled study that consisted of four treatment regimens for 30 months (fluticasone, fluticasone/salmeterol, placebo, or fluticasone the first 6 months followed by placebo for the remaining 24 months of the study). After the end of this 30-month double-blind treatment period, spirometry was repeated once yearly for an additional 5 years(10). After the 30-month trial, ICS treatment was considered ongoing when an individual used ICS for >50% of the total follow-up period(11). Inclusion criteria included an age range from 45 to 75 years old, former or current smokers with at least 10 pack-years, a forced expiratory volume in one second (FEV_1_)/forced vital capacity (FVC) ratio <70%, and FEV_1_ between 30 and 80% of predicted(12). Patients were excluded if they had a history of asthma or when they had used inhaled corticosteroids (ICS) during a period of 6 months prior to inclusion. All patients were clinically stable and allowed to continue using short-acting bronchodilators. Patients were extensively characterised including spirometry, cell differential counts in peripheral blood and induced sputum, and bronchoscopy with bronchial biopsies, which were immunostained with the inflammatory cell markers CD3^+^, CD4^+^, CD8^+^, CD20^+^, CD68^+^, neutrophil elastase, tryptase, and EG2^+^ (13). Methods for RNA isolation from bronchial biopsies, RNA-sequencing, and processing of RNA libraries were described previously(14,15). The local ethics committees approved the study protocol, and all subjects gave their written informed consent.

### Cluster analysis

All analyses were performed in R statistical software (version 4.2.0). The R package ‘ConsensusClusterPlus’ was used to identify clusters based on the 98 genes previously shown to be differentially expressed in the lower airways of patients with COPD compared to non-COPD controls(16). To identify the optimal number of clusters, we calculated a consensus value for each number of clusters, resulting in a value represented by the Cumulative Distribution Functions (CDF). The optimal number of clusters (k) was determined by identifying the point at which the distribution plateaus, indicating maximal stability beyond which further divisions would reflect random rather than true cluster structure(16).

Subsequently, we compared the clinical expression of COPD between the clusters, both cross-sectionally and longitudinally using SPSS 23.0.03 software (SPSS Inc., Chicago, IL, USA). When data were non-normally distributed, a log_2_ transformation was performed. If these variables remained non-normally distributed, original values were used. Differences at baseline and longitudinal clinical features between clusters were compared using independent sample t-tests in case of normal distribution, Mann-Whitney U test for non-normally distributed data and chi-square tests for categorical variables. A linear mixed effects model, corrected for smoking status, ICS treatment (>50% ICS use between 30 months – 7.5 years) and inflammatory cell count, was used to analyse long-term annual FEV_1_ decline (in millilitres per year) between 6 months after the start of the treatment and the end of the follow-up period, as has been done in previous studies(17–19).

To assess how the identified COPD clusters were related to pathobiological differences, we performed whole genome differential gene expression analysis comparing the clusters using a linear model corrected for age, sex, and smoking status (R package *Limma* version *3*.*40*.*6*). Genes were considered to be differentially expressed when their false discovery rate (FDR) was < 0.05. Significantly differentially expressed genes between the COPD clusters were subjected to pathway analysis using G-Profiler and STRING as described previously(20,21).

### Validation of cluster-associated gene expression profiles in independent cohort

To validate our findings, we investigated genes associated with the different COPD clusters in the British Columbia Lung Health Study (BCLHS) with genome-wide gene expression data available from bronchial brushes of smokers (former and current) with and without COPD(8). To this end, we calculated composite scores using Gene-Set Variation Analysis (GSVA) for up-and downregulated genes separately. GSVA scores were then associated with severity of airflow obstruction and rate of lung function decline, using a linear model adjusting for age, sex, pack-years and smoking status.

## Results

Frozen bronchial biopsies with RNA of sufficient quality of 56 patients were available for sequencing. All included patients had cross-sectional and longitudinal data available up to 30 months, and thereafter 31 were followed-up for even longer with annual spirometries for 5 additional years. The mean duration of the follow-up for all patients combined was 5.7±2.6 years. Clinical characteristics of included patients are presented in table 1.

**Table 1.** Clinical features of patients with chronic obstructive pulmonary disease in COPD-associated Airway Gene Expressed 1 (CAGE1) and COPD-associated Airway Gene Expressed 2 (CAGE2)

|  |  | GLUCOLD Cohort<br>N = 56 | CAGE1<br>N = 39 | CAGE2<br>N = 17 |
| --- | --- | --- | --- | --- |
| Sex, male (n, %) |  | 50 (89.3%) | 35 (89.7%) | 15 (88.2%) |
| Age (years) |  | 61.1 (±7.7) | 61.5 (±7.1) | 60.3 (±9.1) |
| BMI (kg/m<sup>2</sup>) |  | 25.3 (±3.6) | 25.2 (±3.1) | 25.5 (±4.6) |
| Current smoking (n, %) |  | 38 (67.9%) | 29 (74.4%) | 9 (52.9%) |
| Smoking load (pack years) |  | 41.4 (31.9 - 54) | 42.0 (31.9 - 55.6) | 36.5 (31.5 - 51.9) |
| Reversibility (≥12% and >200ml improvement) |  | 12 (21.4%) | 8 (20.5%) | 4 (23.5%) |
| GOLD grade (n, %) |  |  |  |  |
|  | GOLD 2 | 51 (91.1%) | 36 (92.3%) | 15 (88.2%) |
|  | GOLD 3 | 5 (8.9%) | 3 (7.7%) | 2 (11.8%) |
| FEV<sub>1</sub> % predicted Post-BD |  | 62.5 (±8.9) | 61.5 (±8.7) | 64.9 (±9.0) |
| FEV<sub>1</sub>/IVC |  | 46.1 (±9.0) | 45.3 (±9.0) | 47.9 (±8.9) |
| RV/TLC |  | 48.2 (±7.8) | 48.8 (±8.3) | 46.9 (±6.6) |
| TLCO % predicted |  | 63.9 (±20.2) | 63.9 (±22.6) | 63.9 (±14.5) |
| PC<sub>20</sub> methacholine (mg/ml) <sup>§</sup> |  | 0.7 (0.2 - 2.4) | 0.7 (0.2 - 2.8) | 0.5 (0.1 - 1.6) |
| PC<sub>20</sub> methacholine (≤8 mg/ml) (n, %) |  | 52 (94.5%) | 36 (94.7%) | 16 (94.1%) |
| CCQ (total score) |  | 1.2 (0.9 - 1.8) | 1.2 (0.8 - 1.8) | 1.4 (0.9 - 1.8) |
| SGRQ (total score) |  | 30 (±14) | 31 (±14) | 29 (±15) |
| Treatment arm: placebo (n, %) |  | 14 (25%) | 8 (20.5%) | 6 (35.3%) |
| Treatment arm: fluticasone/salmeterol (n, %) |  | 16 (29%) | 10 (25.6%) | 6 (35.3%) |
| Treatment arm: fluticasone ≥0 months (n, %) |  | 13 (23%) | 9 (23.1%) | 4 (23.5%) |
| Treatment arm: fluticasone ≥6 months (n, %) |  | 13 (23%) | 12 (30.8%) | 1 (5.9%) |
| ICS use until 30 months (n, %) |  | 42 (75%) | 31 (79.5%) | 11 (64.7%) |
| >50% ICS use of the period 2.5-7.5 years (n, %) |  | 11 (25%) | 9 (29.0%) | 2 (15.4%) |
| Blood | % eosinophils <sup>§</sup> | 2.3 (1.2 - 3.5) | 2.2 (1.3 - 3.2) | 2.7 (1.2 - 4.4) |
|  | % basophils <sup>§</sup> | 0.5 (0.3 - 0.8) | 0.5 (0.3 - 0.7) | 0.4 (0.3 - 1.1) |
|  | % neutrophils | 59.5 (±9.5) | 60.2 (±9.3) | 57.7 (±10.1) |
|  | % monocytes | 8.5 (7.5 - 9.5) | 8.8 (7.5 - 9.5) | 8.3 (7.4 - 10.6) |
|  | % lymphocytes | 28.8 (±8.6) | 28.3 (±7.6) | 29.8 (±10.6) |
| Sputum | % eosinophils <sup>§</sup> | 1.25 (0.3 - 2.3) | 1.3 (0.3 - 2) | 1.0 (0.3 - 2.5) |
|  | % neutrophils | 70.4 (60 - 76.8) | 70 (59.7 - 81.5) | 72 (64 - 75) |
|  | % macrophages | 23.3 (17.9 - 33.6) | 25.2 (15.0 - 34.5) | 21.7 (18.5 - 25.7) |
|  | % lymphocytes | 1.9 (1.3 - 2.9) | 1.8 (1.2 - 2.2) | **2.2 (1.8 - 3.5) #** |
| Biopsy | CD3<sup>+</sup> T-cells (Count / 0.1mm<sup>2</sup>) <sup>§</sup> | 121 (78.9 - 182.1) | 116.5 (64.0 - 169.5) | **164.5 (107.8 - 211.3) #** |
|  | CD4<sup>+</sup> T-cells (Count / 0.1mm<sup>2</sup>) <sup>§</sup> | 57.8 (28.5 - 83.1) | 45 (25.0 - 75.5) | **69.5 (56.5 - 96.5) #** |
|  | CD8<sup>+</sup> T-cells (Count / 0.1mm<sup>2</sup>) <sup>§</sup> | 20.8 (11 - 35.5) | 16 (8.0 - 28.0) | **29.5 (20.3 - 36.5) #** |
|  | NE<sup>+</sup> neutrophils (Count / 0.1mm<sup>2</sup>) <sup>§</sup> | 4 (2.5 - 7.3) | 3 (2.5 - 8.0) | 5 (3.5 - 6.5) |
|  | Tryptase<sup>+</sup> mast cells (Count / 0.1mm<sup>2</sup>) <sup>§</sup> | 28.8 (20.6 - 34.6) | 28.5 (20.5 - 37.0) | 30.5 (23.0 - 32.8) |
|  | CD68<sup>+</sup> macrophages (Count / 0.1mm<sup>2</sup>) | 9.8 (5.1 - 13.4) | 8.5 (5.0 - 13.5) | 12 (9.0 - 13.5) |
|  | EG2<sup>+</sup> eosinophils (Count / 0.1mm<sup>2</sup>) | 1.5 (1 - 4.4) | 1.5 (1 - 4) | 2 (0.8 - 5) |

Description: BMI: body mass index; GOLD: Global Initiative for Chronic Obstructive Lung Disease; FEV<sub>1</sub>: forced expiratory volume in one second; BD: bronchodilator (salbutamol 400mcg); IVC: inspiratory vital capacity; RV: residual volume; TLC: total lung capacity;
TLCO: transfer factor for carbon monoxide; PC<sub>20</sub>: provocative concentration needed for a 20% FEV<sub>1</sub> drop; CCQ: COPD control questionnaire, SGRQ: St. George respiratory questionnaire; ICS: inhaled corticosteroids; NE: neutrophil elastase; §: log2 transformed and presented in geometric mean (IQR). P-values < 0.05 are highlighted in bold and with #.

### Identification of two COPD-associated Airway Gene Expressed clusters, CAGE1 and CAGE2

Cluster analysis showed a model with two clusters to have the lowest CDF value and therefore was chosen for further analysis (figure 1). The first cluster, from now on referred to as COPD-associated Airway Gene Expressed 1 (CAGE1), consisted of 39 COPD patients, and the second one, CAGE2 comprised 17 patients with COPD. The baseline clinical characteristics for CAGE1 and CAGE2 patients are presented in table 1. There were no significant differences between the clusters with respect to sex, age, smoking status, pack years and baseline lung function. The percentage of sputum lymphocytes was higher in CAGE2 compared to CAGE1 patients (2.2% (IQR 1.8 – 3.5) vs. 1.8% (IQR 1.2 – 2.2), p=0.045, figure 2A), as well as CD4^+^ (69.5/0.1mm^2^ (IQR 56.5 – 96.5) vs 45/0.1mm^2^ (IQR 25.0 – 75.5), p=0.04) and CD8^+^ (29.5/0.1mm^2^ (IQR 20.3 – 36.5) vs. 16.0/0.1mm^2^ (IQR 8.0 – 28.0), p=0.01) T-cell counts in biopsies (figure 2B and 2C).

**Figure 1.**
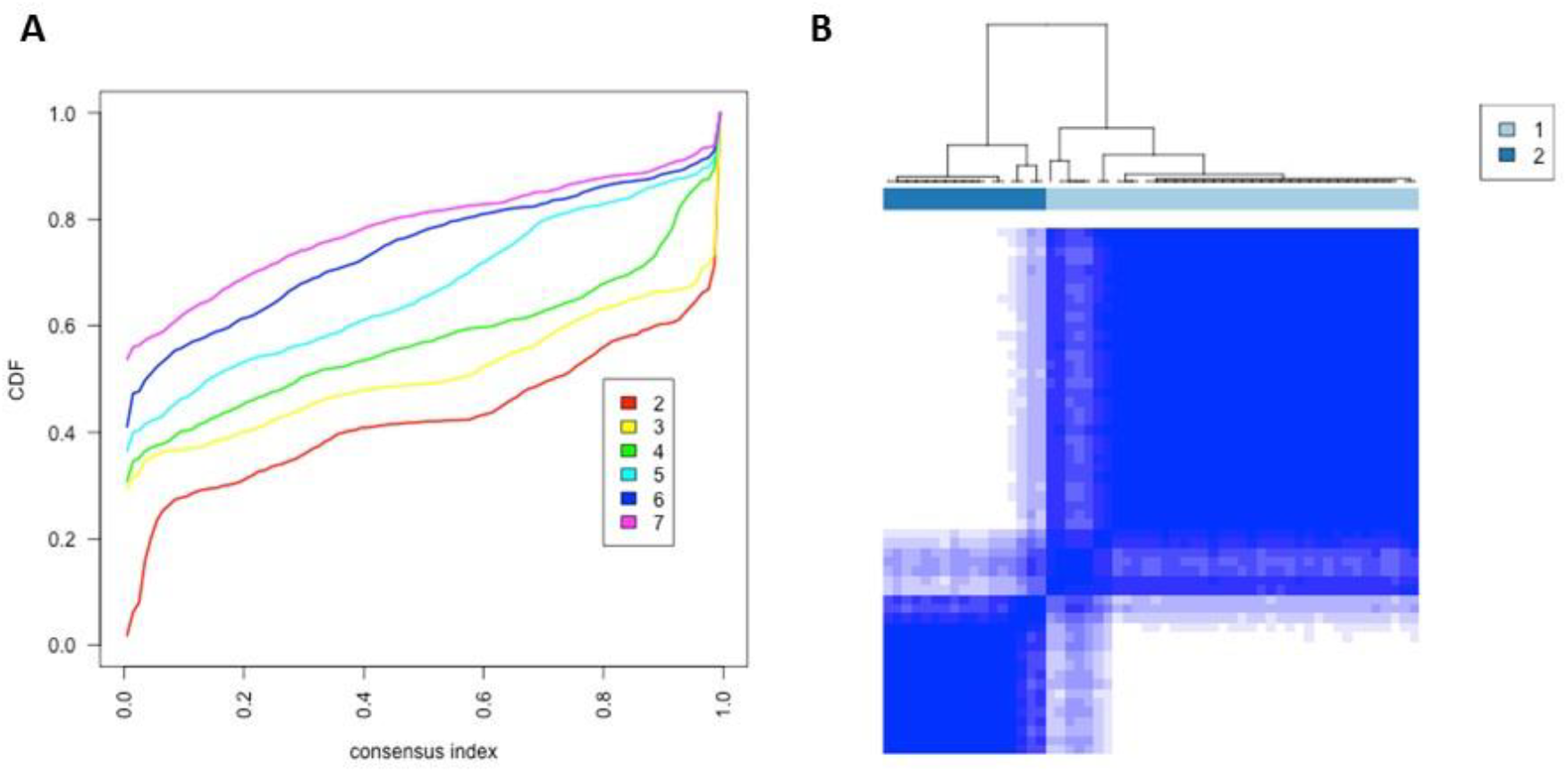
Inter-cluster consensus (A) and clusters’ dimensions (B) Lines in (A) represent the different number of clusters tested. The optimal number of clusters (k) was determined by identifying the point at which the distribution plateaus, indicating maximal stability beyond which further divisions would reflect random rather than true cluster structure. CDF: Cumulative Distribution Functions.

**Figure 2.**
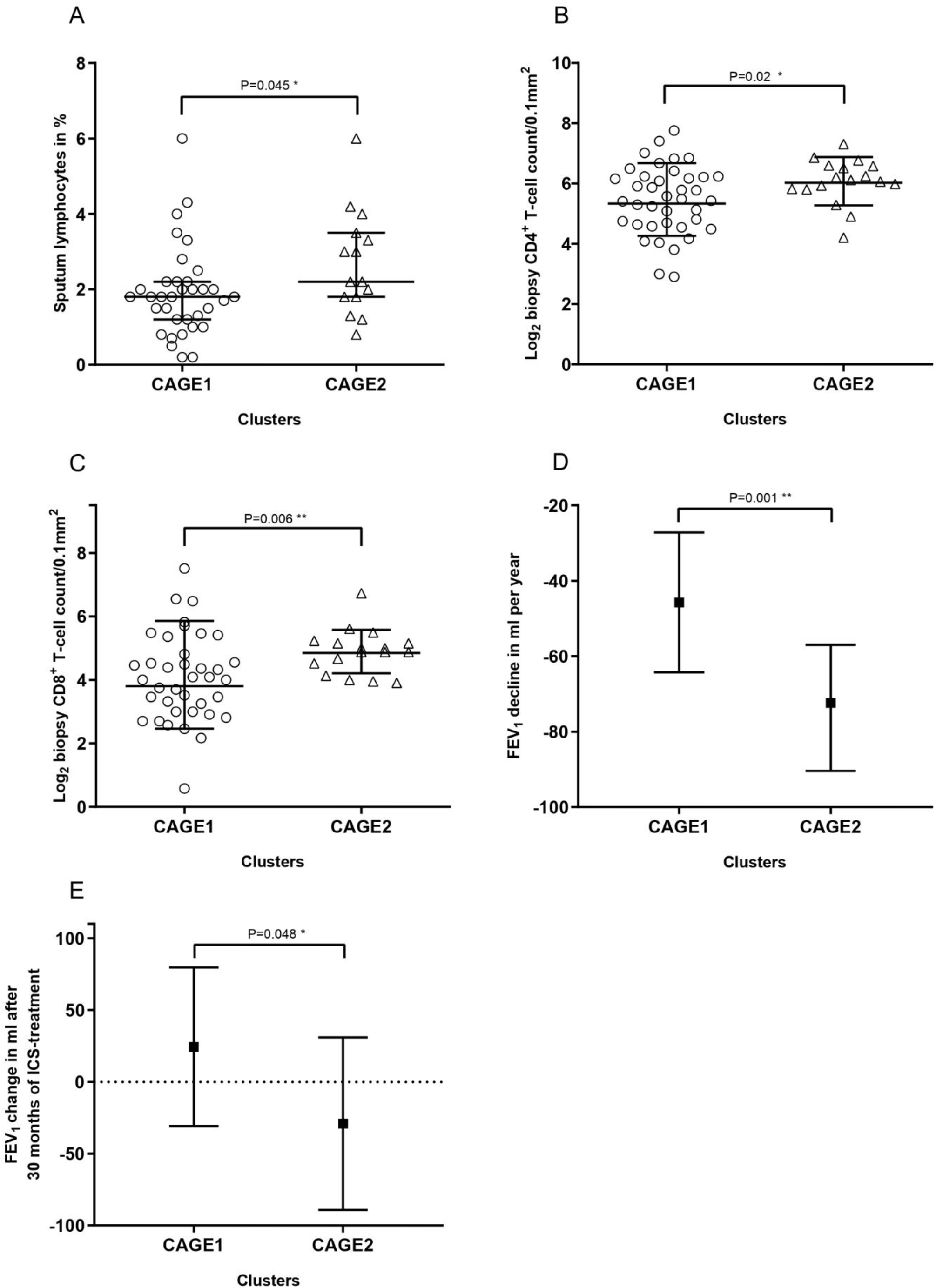
Inflammatory level and lung function parameters in CAGE1 and CAGE2. **2A**: sputum lymphocyte percentage of total amount of cells per cluster at baseline, presented in median with IQR; **2B**: log_2_-transformed biopsy CD4^+^ T-cell count in 0.1mm^2^ per cluster at baseline, presented in geometric mean and SD; **2C**: log_2_-transformed baseline biopsy CD8^+^ T-cell count in 0.1mm^2^ per cluster at baseline, presented in geometric mean and SD; **2D**: annualized FEV_1_ change (ml per year) between 6 months and 7.5 years of follow-up per cluster, corrected for smoking status, ICS treatment and log_2_-transformed CD4^+^ and CD8^+^ T-cell biopsy counts; **2E:** FEV_1_ change (in ml) between 0 and 30 months of ICS treatment per cluster, corrected for smoking status and log_2_-transformed CD4^+^ and CD8^+^ T-cell biopsy counts.

CAGE2 patients experienced a faster decline in their FEV_1_ during the period between 6 months and the end of the follow-up period compared to those in CAGE1 (−69.9 ml/year (95% CI −55.7 to −84.0 ml) vs −44.0 ml/year (95% CI −30.3 to −57.7 ml/year), p=0.002), after correcting for smoking status, ICS treatment and biopsy-derived CD4^+^ and CD8^+^ T-cell counts (figure 2D). In addition, patients in CAGE2 exhibited more severe disease as evidenced by ICS unresponsiveness and a faster lung function decline. After 30 months of treatment with ICS, FEV_1_ improved less in CAGE2 vs CAGE1 patients, the change in FEV_1_ being −29.1ml/year (95% CI −89.2 – 30.9ml) vs +24.4ml/year (95% CI −30.9 – 79.8ml) (p=0.048) respectively (figure 2E), whereas at 6 months there were no differences between the groups (p=0.82, Supplementary figure 1A). Since CD4^+^ and CD8^+^ bronchial biopsy cell counts differed between the clusters, we also analysed their direct association with severity of airflow obstruction and lung function decline. This analysis showed that the number of bronchial biopsy CD4^+^ and CD8^+^ cells were not associated with either baseline FEV_1_% predicted (CD4^+^: Pearson’s R=-0.07, P=0.62; CD8^+^: Pearson’s R=0.13, P=0.33; Supplementary figure 1B and 1C respectively) nor with FEV_1_ decline between 6 months and 7.5 years of follow-up (CD4^+^: Spearman’s R=0.19, P=0.20; CD8^+^: Spearman’s R=0.1, P=0.46; Supplementary figure 1D and 1E respectively).

### Gene expression differences between CAGE1 & CAGE2 and pathway analyses

We identified 200 genes differentially expressed between CAGE1 and CAGE2 with a fold change > |2| and FDR < 0.05. Among these, 186 had a higher and 14 a lower expression in CAGE2 (Supplementary table 1). Pathway analysis using G-profiler showed that genes with higher expression in CAGE2 were enriched for several pro-inflammatory pathways, including antigen binding, humoral immune response mediated by circulating immunoglobin, and B-cell mediated immunity (table 2). No significant enrichment was observed for genes with lower expression in CAGE2. A protein interactome for the identified genes (both with significantly higher and lower expression in CAGE2) was built, to highlight the possible interplay between those genes (figure 3). Two of the previously identified COPD genes, *LTF* and *C6orf58*, are representing important connections in the middle of the network (red nodes identifying the top 20 genes with higher expression in CAGE2), with *LTF* being among the genes with the highest number of connections (purple nodes identifying the top 5 most connected genes). The genes with lower expression in CAGE2 (blue nodes identifying the top 10) were also involved in the same pathway as the genes with higher expression in CAGE2 (i.e. *SERPINB2*). Additionally, some genes (i.e. *CYP3A5*) were not part of the main network but instead involved in smaller and independent networks.

**Table 2.** Top 10 pathways identified in the higher expression genes associated with CAGE2 cluster.

| Rank | Term name | Term ID | Term genes (N) | Query genes (N) | Common genes (N) | Corrected P |
| --- | --- | --- | --- | --- | --- | --- |
| 1. | Antigen binding | G0:0003823 | 189 | 164 | 19 | 6.07 x 10<sup>-12</sup> |
| 2. | Complement activation, classical pathway | G0:0006958 | 133 | 164 | 15 | 1.08 x 10<sup>-9</sup> |
| 3. | Immune response-activating signal transduction | G0:0002757 | 576 | 164 | 27 | 1.10 x 10<sup>-9</sup> |
| 4. | Activation of immune response | G0:0002253 | 638 | 164 | 28 | 1.91 x 10<sup>-9</sup> |
| 5. | Immune response-regulating signalling pathway | G0:0002764 | 605 | 164 | 27 | 3.48 x 10<sup>-9</sup> |
| 6. | Humoral immune response mediated by circulating immunoglobulin | G0:0002455 | 144 | 164 | 15 | 3.50 x 10<sup>-9</sup> |
| 7. | Immunoglobulin mediated immune response | G0:0016064 | 205 | 164 | 17 | 4.51 x 10<sup>-9</sup> |
| 8. | B cell mediated immunity | G0:0019724 | 207 | 164 | 17 | 5.28 x 10<sup>-9</sup> |
| 9. | Positive regulation of immune response | G0:0050778 | 775 | 164 | 30 | 6.41 x 10<sup>-9</sup> |
| 10. | Adaptive immune response | G0:0002250 | 488 | 164 | 24 | 9.10 x 10<sup>-9</sup> |

**Figure 3.**
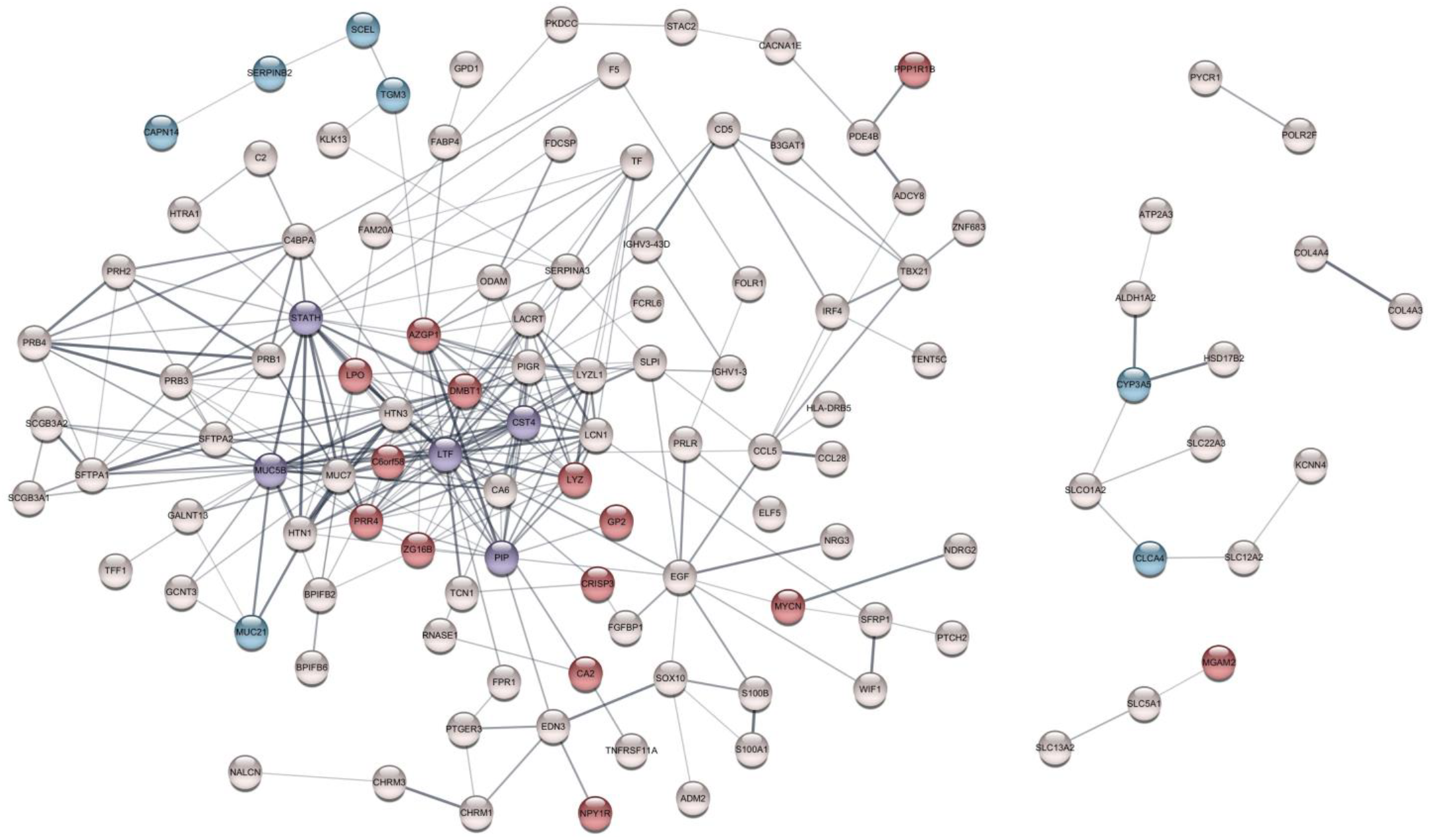
Interactome network of CAGE2-associated differentially expressed genes. Blue-filled nodes represent the top 10 genes with lower expression in CAGE2. Red-filled nodes represent the top 20 genes with higher expression in CAGE2. Purple-filled nodes represent the top 5 most connected genes in the network. Lines between nodes represent an interaction.

### Validation of the CAGE2 COPD endotype in relation to disease severity

We used GSVA composite scores of genes with higher or lower expression in CAGE2 to assess their association with severity of COPD in the 87 patients with COPD included in the British Columbia Lung Health Study. Their clinical characteristics are presented in supplementary table 2. We found the GSVA score of genes with higher expression in CAGE2 to be associated with more severe airflow obstruction as reflected by a lower FEV_1_/FVC (t-value=-2.74, p=7.44 × 10^−3^), with a trend for a lower baseline FEV_1_ % predicted (t-value=-1.70, p=0.09, table 3). No significant associations with baseline lung function measurements were observed for the GSVA score of genes with lower expression in CAGE2 and neither GSVA scores associated with lung function decline (GSVA for genes with higher expression in CAGE2: t-value=-0.35, p=0.73, and for genes with lower expression in CAGE2: t-value=0.15, P=0.88, table 3).

**Table 3.** Validation of CAGE2 signature in patients with chronic obstructive pulmonary disease of the British Columbia Lung Health Study (n=87).

|  | T<br>GSVA of higher expression<br>genes in CAGE2 vs. CAGE1 | P | T<br>GSVA of lower expression<br>genes in CAGE2 vs. CAGE1 | P |
| --- | --- | --- | --- | --- |
| ΔFEV<sub>1</sub> (ml per year) | -0.35 | 0.73 | 0.15 | 0.88 |
| Baseline FEV<sub>1</sub> % predicted (%) | -1.70 | 0.09 | -1.42 | 0.16 |
| FEV<sub>1</sub>/FVC ratio (%) | **-2.74** | **7.44 x 10<sup>-3</sup>** | -0.04 | 0.97 |

Description: ΔFEV<sub>1</sub>: change in forced expiratory volume in 1 second; FVC: forced vital capacity; GSVA: gene-set enrichment analysis; CAGE: COPD-associated Airway Gene Expressed. Corrected for sex, age, pack-years, and smoking status.

## Discussion

In the current study, we identified two COPD endotypes, termed CAGE1 and CAGE2. Patients with CAGE2 COPD exhibited a different inflammatory profile in their sputum and airways along with more severe disease as reflected by ICS unresponsiveness and faster lung function decline. Pathway analysis unveiled a unique pathobiology in CAGE2, marked by more activation of T- and B-cell immune responses. Our findings underscore the utility of endotyping in delineating clinically relevant COPD subgroups with direct ties to their underlying pathobiology, offering promising prospects for future research on a larger scale.

We found CAGE2 to be associated with more severe disease and a distinct inflammatory profile characterized by elevated sputum lymphocytes and higher CD4^+^ and CD8^+^ T-cell counts in the airway wall. Our findings are in line with previous studies showing bronchial CD4^+^ and CD8^+^ T-cells to be associated with more severe lung function impairment in COPD (22–24). Higher percentage of airways containing CD4^+^ and CD8^+^ T-cells in surgically resected lung tissue have also been shown to be associated with more severe COPD GOLD stage (22). Although higher CD4^+^ and CD8^+^ cell counts are related with more severe COPD and worse outcome (25–27), it is important to note that we observed a faster lung function decline in CAGE2 COPD, even when adjusting for baseline bronchial CD8^+^ and CD4^+^ T-cell counts. Therefore, other factors, independent of CD4^+^ and CD8^+^ inflammatory cells, are likely involved in this rapid FEV_1_ decline in CAGE2 patients.

Of particular interest are four genes in the interactome of CAGE2-associated genes: Lactotransferrin (*LTF*) and Chromosome 6 Open Reading Frame 58 (*C6orf58*) with higher, and Cytochrome P450 Family 3 Subfamily A Member 5 (*CYP3A5*) and Serpin Family B Member 2 (*SERPINB2*) with lower expression in CAGE2. LTF is a globular glycoprotein widely present in secretory body fluids, which has both direct and indirect antimicrobial effects (28), and was previously found to be elevated in non-typeable Haemophilus influenzae and seasonal influenza A virus infections (29). Our findings of higher LTF may suggest a role for bacterial colonization and/or more frequent infections contributing to more severe disease in CAGE2 COPD. Although the exact function of the C6orf58 gene, apart from being involved in liver development in zebra fish, is unknown (30,31), C6orf58 has been previously identified with higher expression in the sputum supernatant of patients with COPD compared to non-COPD controls (31). This finding combined with our results suggest a possible role for C6orf58 in COPD which deserves more study. The gene *CYP3A5* with lower expression in CAGE2 is a well-known member of the cytochrome P450 superfamily of enzymes, which metabolize (inhaled) toxicants (such as tobacco smoke)(32,33). It could be speculated that a lower *CYP3A5* expression may impair detoxification responses, thereby increasing the susceptibility to develop COPD (32). In line with this hypothesis, and our findings, a *CYP3A5* gene polymorphism leading to lower expression of the gene in macrophages has been shown to be associated with a faster FEV_1_ decline in current smokers (33). Finally, the *SERPINB2* gene had lower expression in CAGE2. *SERPINB2* enables transcription of the plasminogen activator inhibitor 2, a coagulation factor abundantly present in epithelial cells, monocytes, and macrophages(34). It has been previously observed that a higher epithelial *SERPINB2* expression in respiratory epithelial cells was associated with lower FEV_1_/FVC, FEV_1_ % predicted, and worse disease severity in asthmatic patients (35). Another interesting observation in this context is that lower expression of this gene is associated with ICS unresponsiveness in asthma, which is in line with our findings in CAGE2 (36).

To date, only a limited number of studies have conducted clustering on COPD patients using gene expression signatures. Notably, the ECLIPSE study analysed genome-wide blood gene expression from 229 former smokers (7), identifying four distinct clinical subtypes of COPD, which were successfully reproduced in an independent cohort. The four groups were characterized by differences in baseline FEV_1_/FVC, FEV_1_, emphysema and symptom severity. While blood sampling offers easier clinical application compared to taking bronchial biopsies, our current analyses indicate that the CAGE2 gene expression cluster identified at baseline is more strongly linked to prospective lung function decline rather than existing lung damage, thus yielding more prognostic value. For a biomarker that can predict future risk of rapid lung function loss, a more invasive diagnostic method such as bronchoscopy may be warranted, as it provides a more comprehensive representation of the activity of all the cells involved in the disease process, whereas analysis of white blood cells is limited to (off-site) inflammatory cells alone.

In the validation cohort, we observed a strong association between higher expression of genes in CAGE2 and more severe airflow obstruction, as indicated by the GSVA score. We did not find the GSVA composite score of genes with higher expression in CAGE2 to be associated with lung function decline in the validation cohort. A possible explanation for the lack of an association with lung function decline could be the fact that more severe COPD patients have less room to deteriorate further in terms of lung function, as has been observed in many studies (37,38).

Our study is the first to apply a clustering method on gene expression data derived from bronchial biopsies in patients with COPD. A possible limitation is the low number of patients included in our study and the lack of more independent cohorts to replicate our findings, as studies that performed a detailed clinical characterization including bronchoscopy combined with long-term follow-up in COPD are scarce. In addition, our study was confined to endotyping based solely on gene expression, which although of value, may not always align with protein expression that ultimately influences biological effects. Nevertheless, our findings are of interest and pave the way for future large-scale studies aiming to endotype COPD, preferably not only based on gene expression, but including other relevant data layers, such as the proteome.

In conclusion, our cluster analysis based on bronchial gene expression profiles identified two COPD endotypes, CAGE1 and CAGE2. We show that CAGE2 COPD endotype is associated with more severe disease with ICS unresponsiveness and more rapid lung function decline. In addition, we show a different pathobiology in CAGE2 COPD, with more activation of T- and B-cell immune responses and differential expression of several key genes that may contribute to the more severe disease observed in this endotype. These findings highlight the utility of endotyping to identify clinically relevant subgroups of COPD and elucidate their underlying pathobiology.

## Data Availability

All data produced in the present study are available upon reasonable request to the authors.

## Conflict of interest

A. Spira is an employee of Johnson and Johnson.

## Notes

**Funding**: The study was sponsored by the Dutch Ministry of Health via the Public Private Funding Program (PPP), GlaxoSmithKline (The Netherlands), the University Medical Centre Groningen and the Leiden University Medical Centre. Rui Marçalo is supported by Fundação para a Ciência e Tecnologia through the grant 10.54499/UI/BD/151337/2021.

### Clinical Trial

NCT00158847

### Funding Statement

The study was sponsored by the Dutch Ministry of Health via the Public Private Funding Program (PPP), GlaxoSmithKline (The Netherlands), the University Medical Centre Groningen and the Leiden University Medical Centre. Rui Marcalo is supported by Fundacao para a Ciencia e Tecnologia through the grant 10.54499/UI/BD/151337/2021.

### Author Declarations

Ethics committee of Groningen University Medical Center gave ethical approval for this work. Ethics committee of Leiden University Medical Center gave ethical approval for this work.

