## Supplementary figure 1 for "Bronchial gene expression clustering in COPD identifies a subgroup of patients with higher level of bronchial T-cells and accelerated lung function decline"

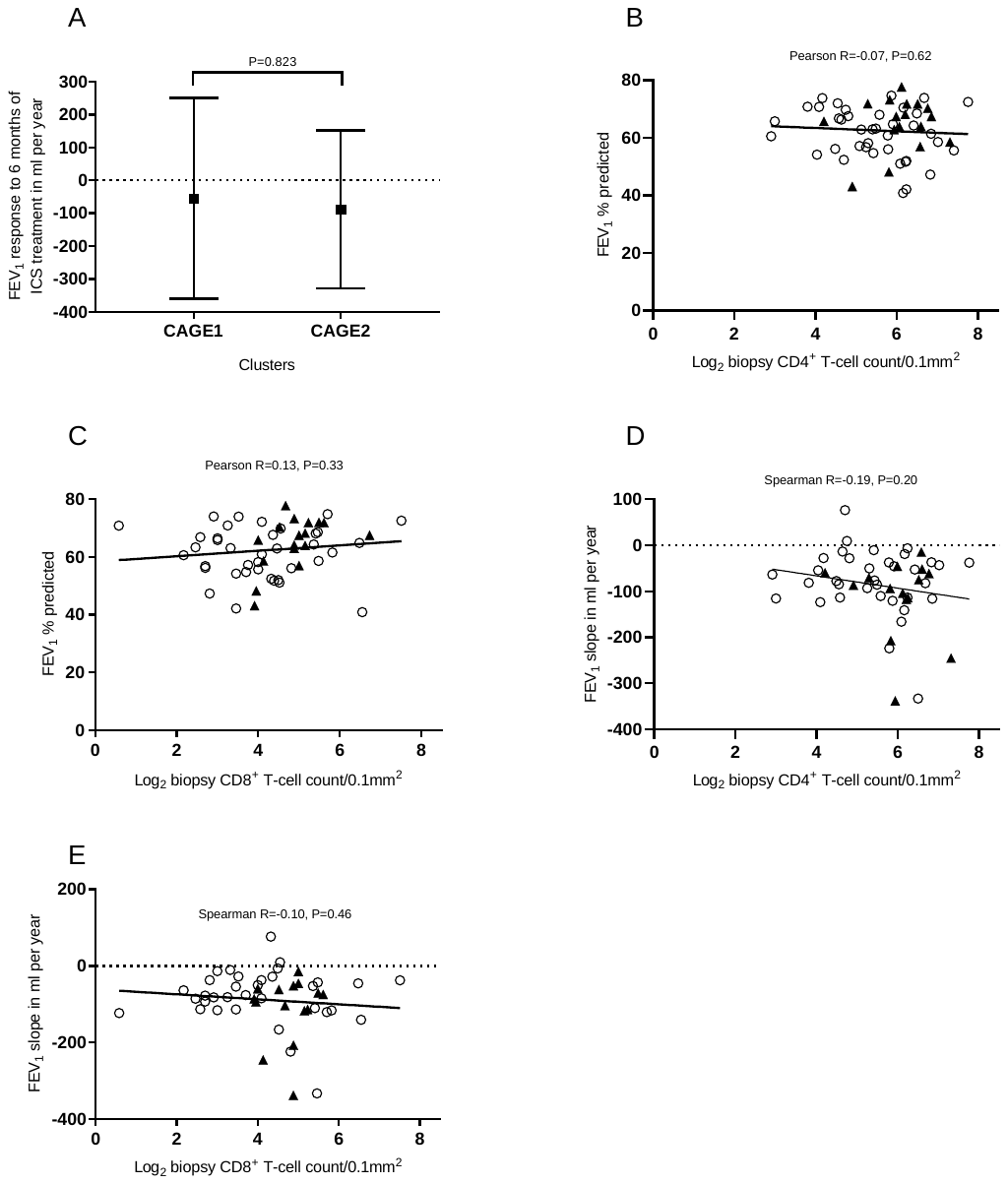

**Supplementary figure 1**. **A**: FEV_1_ change in ml between 0 and 6 months of ICS treatment per cluster, corrected for smoking status and log_2_-transformed CD4^+^ and CD8^+^ T-cell biopsy count variance from geometric mean. **B**: correlation between baseline FEV_1_ % predicted and baseline log_2_-transformed biopsy CD4^+^ T-cell count; **C**: correlation between baseline FEV_1_ % predicted and baseline log_2_-transformed biopsy CD8^+^ T-cell count; **D**: correlation between individual FEV_1_ change in ml per year between 6 months - 7.5 years and baseline log_2_-transformed biopsy CD4^+^ T-cell count; **E**: correlation between individual FEV_1_ change in ml per year between 6 months - 7.5 years and baseline log_2_-transformed biopsy CD8^+^ T-cell count. White circles correspond to patients from CAGE1 cluster, whereas black triangles correspond to patients from CAGE2 cluster.
