## Supplementary table 1 for "Bronchial gene expression clustering in COPD identifies a subgroup of patients with higher level of bronchial T-cells and accelerated lung function decline"

**Supplementary table 1**: Significantly up- and down-regulated genes associated with CAGE2 cluster compared to CAGE1.

| **Symbol** | **Log_2_-fold change** | **False Discovery Rate** |
| --- | --- | --- |
| NXPE2 | 2.58 | 2.05E-24 |
| LYZ | 2.51 | 7.58E-21 |
| LTF | 2.35 | 8.37E-18 |
| AZGP1 | 2.32 | 8.81E-22 |
| DMBT1 | 2.23 | 5.26E-16 |
| ZG16B | 2.22 | 5.07E-19 |
| MGAM2 | 2.21 | 8.00E-17 |
| PPP1R1B | 2.18 | 1.19E-18 |
| CRISP3 | 2.17 | 2.91E-15 |
| CA2 | 2.15 | 5.75E-18 |
| PRR4 | 2.15 | 1.09E-17 |
| PIP | 2.15 | 7.90E-15 |
| CCDC129 | 2.15 | 2.68E-15 |
| GP2 | 2.07 | 3.20E-14 |
| TTYH1 | 2.05 | 1.27E-14 |
| LINC02009 | 2.03 | 1.09E-13 |
| C6orf58 | 2.03 | 3.26E-13 |
| MYCN | 2.02 | 8.00E-17 |
| LPO | 2.00 | 9.81E-13 |
| NPY1R | 1.98 | 4.08E-15 |
| SLC13A2 | 1.97 | 1.14E-17 |
| AL603764.1 | 1.97 | 1.22E-12 |
| TF | 1.96 | 1.67E-13 |
| SLCO1A2 | 1.96 | 1.70E-12 |
| BPIFB2 | 1.95 | 9.81E-13 |
| AC078923.1 | 1.94 | 3.05E-12 |
| NRG3 | 1.94 | 1.72E-12 |
| CHRM1 | 1.94 | 9.66E-13 |
| ALDH1A2 | 1.94 | 2.30E-15 |
| PADI2 | 1.92 | 1.87E-20 |
| ALDH1L2 | 1.90 | 5.07E-19 |
| FDCSP | 1.88 | 2.97E-12 |
| LINC01829 | 1.86 | 4.25E-12 |
| PRLR | 1.84 | 6.63E-15 |
| SETP21 | 1.83 | 3.80E-12 |
| S100A1 | 1.81 | 7.76E-11 |
| RTN1 | 1.78 | 4.85E-15 |
| EGF | 1.77 | 3.97E-16 |
| MUC5B | 1.76 | 1.78E-10 |
| SERPINA3 | 1.75 | 6.94E-10 |
| AL603764.2 | 1.73 | 1.41E-09 |
| PRH2 | 1.72 | 1.16E-09 |
| ADCY8 | 1.72 | 1.53E-09 |
| ODAM | 1.71 | 1.60E-09 |
| AC061979.1 | 1.69 | 1.82E-09 |
| DLGAP1-AS5 | 1.67 | 1.89E-10 |
| CGREF1 | 1.67 | 4.68E-10 |
| OXGR1 | 1.65 | 6.51E-11 |
| LINC02188 | 1.65 | 1.70E-11 |
| SLC5A1 | 1.63 | 5.20E-14 |
| PKDCC | 1.60 | 1.46E-10 |
| LINC02473 | 1.57 | 5.44E-11 |
| IGKV2-24 | 1.56 | 7.22E-08 |
| MTCO3P12 | 1.55 | 1.16E-07 |
| PART1 | 1.55 | 5.37E-14 |
| PRB3 | 1.54 | 1.20E-07 |
| GPD1 | 1.53 | 5.87E-13 |
| FOLR1 | 1.52 | 1.20E-11 |
| ASTN1 | 1.52 | 4.52E-08 |
| AC087525.1 | 1.51 | 3.92E-12 |
| TFF1 | 1.50 | 5.73E-08 |
| STATH | 1.49 | 4.96E-07 |
| STAC2 | 1.49 | 1.09E-07 |
| SOX10 | 1.48 | 9.64E-08 |
| KCNN4 | 1.46 | 2.43E-11 |
| SLPI | 1.46 | 1.80E-18 |
| FAM20A | 1.46 | 2.69E-09 |
| S100B | 1.45 | 2.71E-08 |
| GALNT13 | 1.45 | 1.94E-07 |
| EDN3 | 1.45 | 7.70E-07 |
| SYT7 | 1.45 | 2.67E-10 |
| B3GAT1 | 1.44 | 4.50E-08 |
| IGHV6-1 | 1.43 | 1.60E-06 |
| AC009041.2 | 1.41 | 6.10E-08 |
| CDR1 | 1.41 | 4.25E-08 |
| FBN3 | 1.40 | 9.90E-07 |
| TCN1 | 1.40 | 1.37E-08 |
| ELF5 | 1.40 | 4.33E-10 |
| IGKV3D-20 | 1.38 | 4.13E-06 |
| RNASE1 | 1.37 | 1.81E-12 |
| SYT13 | 1.37 | 1.18E-08 |
| SOX9-AS1 | 1.37 | 2.85E-09 |
| GLYATL2 | 1.36 | 4.28E-08 |
| AFF3 | 1.36 | 6.45E-09 |
| BHLHA15 | 1.36 | 3.51E-06 |
| PSD2 | 1.35 | 8.85E-09 |
| LINC01482 | 1.35 | 2.64E-08 |
| AC138517.2 | 1.35 | 2.44E-07 |
| PIK3AP1 | 1.34 | 1.14E-11 |
| IGHM | 1.33 | 1.16E-05 |
| SCGB3A2 | 1.32 | 2.40E-06 |
| ATP2A3 | 1.31 | 9.01E-10 |
| SFTPA2 | 1.31 | 1.67E-06 |
| LINC02095 | 1.31 | 1.66E-05 |
| C2 | 1.29 | 7.65E-13 |
| IGLV1-36 | 1.29 | 2.04E-05 |
| SLC12A2 | 1.29 | 5.01E-14 |
| INPP5J | 1.28 | 1.16E-08 |
| F5 | 1.27 | 1.80E-06 |
| PRB4 | 1.27 | 1.54E-05 |
| IGHV3-73 | 1.26 | 4.44E-05 |
| SFRP1 | 1.25 | 9.22E-08 |
| CCL28 | 1.24 | 5.55E-09 |
| LINC01239 | 1.24 | 1.54E-06 |
| SCGB3A1 | 1.23 | 2.13E-05 |
| ZNF683 | 1.22 | 2.73E-06 |
| BPIFB6 | 1.22 | 8.64E-05 |
| FABP4 | 1.21 | 1.44E-05 |
| KANK4 | 1.20 | 1.46E-05 |
| DACH2 | 1.20 | 1.62E-05 |
| KDELC1P1 | 1.19 | 3.67E-08 |
| SLC22A3 | 1.19 | 5.51E-06 |
| PTCH2 | 1.19 | 1.33E-06 |
| PRB1 | 1.19 | 8.28E-05 |
| TESC | 1.18 | 1.03E-05 |
| SLC28A3 | 1.18 | 5.73E-08 |
| SNPH | 1.18 | 4.05E-08 |
| AL139383.1 | 1.17 | 3.95E-08 |
| POLR2F | 1.17 | 1.40E-06 |
| SSTR5-AS1 | 1.16 | 1.36E-04 |
| NKX3-1 | 1.16 | 6.82E-06 |
| CD5 | 1.15 | 2.73E-06 |
| AP003559.1 | 1.15 | 7.92E-05 |
| IGHV4-4 | 1.15 | 2.38E-04 |
| SMOC1 | 1.15 | 6.96E-06 |
| MAEL | 1.15 | 2.58E-04 |
| CHRM3 | 1.14 | 3.51E-06 |
| GCNT3 | 1.14 | 3.56E-05 |
| CACNA1E | 1.14 | 1.59E-04 |
| IGLV3-19 | 1.13 | 3.22E-04 |
| IGKV3-20 | 1.13 | 2.88E-04 |
| AL445437.1 | 1.13 | 1.99E-05 |
| KLHDC7B | 1.13 | 1.10E-04 |
| IGHV2-5 | 1.12 | 3.66E-04 |
| HLA-DRB5 | 1.11 | 6.12E-05 |
| FCRL6 | 1.11 | 2.67E-05 |
| HSD17B2 | 1.11 | 3.00E-06 |
| TNFRSF11A | 1.11 | 2.21E-10 |
| SMPDL3B | 1.11 | 8.42E-07 |
| NALCN | 1.10 | 9.95E-06 |
| IGKV1-27 | 1.10 | 4.69E-04 |
| ADGRG2 | 1.10 | 6.47E-06 |
| IGHV1-3 | 1.10 | 4.66E-04 |
| GFPT2 | 1.10 | 2.10E-06 |
| PTGER3 | 1.08 | 3.26E-05 |
| IGHV1OR15-9 | 1.08 | 6.92E-04 |
| AC064875.1 | 1.07 | 8.00E-05 |
| ZBP1 | 1.07 | 9.60E-05 |
| IGLV1-44 | 1.07 | 7.57E-04 |
| COL4A3 | 1.07 | 5.21E-05 |
| CCL5 | 1.06 | 1.17E-05 |
| HTRA1 | 1.06 | 3.30E-10 |
| KCNH8 | 1.06 | 3.00E-06 |
| NDRG2 | 1.06 | 8.17E-13 |
| IRF4 | 1.06 | 1.79E-04 |
| PYCR1 | 1.06 | 1.43E-05 |
| LINC01554 | 1.06 | 1.35E-05 |
| RASAL1 | 1.06 | 3.70E-06 |
| ADM2 | 1.06 | 4.89E-05 |
| FAM46C | 1.05 | 9.64E-08 |
| FRMD5 | 1.05 | 1.07E-04 |
| LINC02344 | 1.05 | 3.62E-06 |
| GRB14 | 1.05 | 1.83E-04 |
| CDH12 | 1.05 | 2.00E-05 |
| AC233755.2 | 1.05 | 1.00E-03 |
| AP004608.1 | 1.04 | 4.66E-04 |
| P2RX5 | 1.04 | 3.46E-04 |
| IGKV2-30 | 1.03 | 1.22E-03 |
| CA6 | 1.03 | 1.25E-03 |
| TBC1D27 | 1.03 | 9.14E-04 |
| TBX21 | 1.03 | 2.21E-04 |
| IGHG3 | 1.03 | 1.25E-03 |
| IGHV3-13 | 1.02 | 1.15E-03 |
| NXF3 | 1.02 | 2.31E-04 |
| DEPTOR | 1.02 | 2.89E-11 |
| RAP1GAP2 | 1.02 | 3.09E-08 |
| IGKV1-9 | 1.02 | 1.45E-03 |
| WIF1 | 1.02 | 1.49E-03 |
| CLDN22 | 1.01 | 1.61E-04 |
| AL512306.3 | 1.01 | 9.84E-05 |
| IGLV1-40 | 1.01 | 1.62E-03 |
| FPR1 | 1.00 | 7.74E-04 |
| NCMAP | 1.00 | 5.49E-08 |
| PDE4B | 1.00 | 1.23E-05 |
| MEG9 | 1.00 | 6.53E-04 |
| COL4A4 | 1.00 | 3.93E-05 |
| SCEL | -1.27 | 2.57E-05 |
| CLCA4 | -1.18 | 8.80E-05 |
| CAPN14 | -1.15 | 2.51E-04 |
| TGM3 | -1.13 | 3.24E-04 |
| NMRAL2P | -1.13 | 1.07E-04 |
| SERPINB2 | -1.11 | 4.12E-04 |
| A2ML1 | -1.11 | 4.24E-04 |
| MUC21 | -1.10 | 3.88E-04 |
| HS6ST2 | -1.07 | 1.46E-06 |
| CYP3A5 | -1.05 | 4.59E-04 |
| SLC7A11 | -1.05 | 3.27E-04 |
| FGFBP1 | -1.04 | 1.02E-03 |
| KLK13 | -1.02 | 1.46E-03 |
| LYPD3 | -1.01 | 1.44E-03 |
