## Supplementary table 2 for "Bronchial gene expression clustering in COPD identifies a subgroup of patients with higher level of bronchial T-cells and accelerated lung function decline"

**Supplementary table 2**: baseline demographics of the validation cohort.

|  | **BCLHS** | **GLUCOLD** |
| --- | --- | --- |
| N | 87 | 56 |
| Mean age | 65 ±6 | 61.1 ± 7.7 |
| Mean pack years | 51 ±25 | 41.4 [31.9 – 54] |
| Current smokers (n, %) | 30 (34.5%) | 38 (67.9%) |
| Sex, male (n, %) | 35 (40.2%) | 50 (89.3%) |
| Inhaled medications (n, %) | 23 (26.4%) | Placebo: 14 (25.0%)  Fluticasone/salmeterol: 16 (28.6%)  Fluticasone 6 months: 13 (23.2%)  Fluticasone 30 months: 13 (23.2%) |
| Baseline FEV_1_ % predicted | 62.5 ± 8.9 | 62.5 ± 8.9 |
| ΔFEV_1_ (ml/year) | -40.0 ± 50.0 | -66.1 [-59.6 - -72.7] |
| Follow up time (years) | 4 | 5.7 ±2.6 |

FEV_1_: forced expiratory volume in 1 second.
